# The absence of islet autoantibodies in clinically diagnosed older-adult onset type 1 diabetes suggests an alternative pathology, advocating for routine testing in this age group

**DOI:** 10.1101/2021.03.22.21252507

**Authors:** Nicholas J Thomas, Helen C Walkey, Akaal Kaur, Shivani Misra, Nick S Oliver, Kevin Colclough, Michael N Weedon, Desmond G Johnston, Andrew T Hattersley, Kashyap A Patel

**Affiliations:** Institute of Biomedical and Clinical Science, University of Exeter Medical School, Exeter, UK; Department of Diabetes and Endocrinology, Royal Devon and Exeter NHS Foundation Trust, Exeter, UK, Exeter, UK; Faculty of Medicine, Imperial College, London, UK

**Author notes:** **Corresponding author** Kashyap A Patel, Institute of Biomedical and Clinical Science & NIHR Exeter Clinical Research Facility, University of Exeter, College of Medicine and Health, Exeter, UK.

## Abstract

**Objective:** Islet autoantibodies at diagnosis are not well studied in older-adult onset (>30years) type 1 diabetes due to difficulties of accurate diagnosis. We used a type 1 diabetes genetic risk score (T1DGRS) to identify type 1 diabetes aiming to evaluate the prevalence and pattern of autoantibodies in older-adult onset type 1 diabetes.

**Methods:** We used a 30 variant T1DGRS in 1866 white-European individuals to genetically confirm a clinical diagnosis of new onset type 1 diabetes. We then assessed the prevalence and pattern of GADA, IA2A and ZnT8A within genetically consistent type 1 diabetes across three age groups (<18years (n=702), 18-30years (n=524) and >30years (n=588)).

**Findings:** In autoantibody positive cases T1DGRS was consistent with 100% type 1 diabetes in each age group. Conversely in autoantibody negative cases, T1DGRS was consistent with 93%(56/60) of <18years, 55%(37/67) of 18-30years and just 23%(34/151) of >30years having type 1 diabetes. Restricting analysis to genetically consistent type 1 diabetes showed similar proportions of positive autoantibodies across age groups (92% <18years, 92% 18-30years, 93% >30years)[p=0.87]. GADA was the most common autoantibody in older-adult onset type 1 diabetes, identifying 95% of autoantibody positive cases versus 72% in those <18years.

**Interpretation:** Older adult-onset type 1 diabetes has identical rates but different patterns of positive autoantibodies to childhood onset. In clinically suspected type 1 diabetes in older-adults, absence of autoantibodies strongly suggests non-autoimmune diabetes. Our findings suggest the need to change guidelines from measuring islet autoantibodies where there is diagnostic uncertainty to measuring at least GADA in all suspected adult type 1 diabetes cases.

## Introduction

Type 1 diabetes is an autoimmune disease characterized by the presence of circulating islet autoantibodies (glutamate decarboxylase (GADA), islet antigen-2 (IA2A), zinc transporter 8 (ZnT8A) and insulin autoantibodies (IAA)) that can be used to support the diagnosis in routine clinical practice. Controversy exists over the role of routine testing of islet autoantibodies in clinically suspected type 1 diabetes cases. In children absence of islet autoantibodies increases the likelihood of monogenic diabetes (1) but this still remains unlikely with the vast majority of cases without islet autoantibodies having type 1 diabetes (2–5). That islet autoantibodies can be absent at diagnosis in people with type 1 diabetes has likely formed the basis for some clinical guidelines advising against routine testing of islet autoantibodies in suspected type 1 diabetes cases, regardless of age at onset (6–8).

The main difficulties in accurate type 1 diabetes diagnosis are predominantly in older-adults (diagnosed over 30 years of age) not in children. The relative rarity and overlapping clinical features of type 1 diabetes compared to type 2 diabetes after 30 years of age make it difficult to correctly classify type 1 diabetes at diagnosis (9–12). In older adults it is not known if the recommendation of not routinely testing islet autoantibodies in clinically suspected type 1 diabetes cases is still applicable. This is particularly important given the frequent observation that islet autoantibody negative type 1 diabetes is far more common with increasing age of diagnosis (2–4, 13). In older adults ~30% of all clinically diagnosed type 1 diabetes cases are islet autoantibody negative (4, 13–16) versus ~10% in children (1–4). The cause of this increase in negative islet autoantibodies in older-adult onset clinically diagnosed type 1 diabetes is unknown. It could represent an underlying difference in the pathophysiology of type 1 diabetes with increasing age of diagnosis (17, 18). Or could suggest the inclusion of people with non-type 1 diabetes, misclassified as type 1 diabetes thereby increasing the proportion of observed negative islet autoantibody cases. We and others have shown that misdiagnosis of older-adult onset type 1 diabetes is common (12, 19, 20). A high frequency of non-type 1 diabetes contributing to islet antibody negative clinically diagnosed adult onset type 1 diabetes would support routine testing of islet autoantibodies in this age group.

Understanding the frequency of islet autoantibodies in older-adult onset type 1 diabetes at diagnosis requires an independent method of confirming type 1 diabetes in clinically suspected cases without relying on islet autoantibody measurement itself. C-peptide deficiency in longstanding disease allows a gold standard biological definition of type 1 diabetes but is not useful close to diagnosis (21, 22). We have recently shown that a method based on distributions of genetic susceptibility to type 1 diabetes in people with diabetes, assessed using genetic risk scores, can be used determine the prevalence of type 1 and non-type 1 diabetes in a cohort (23, 24). This approach, which can be used at any time after diagnosis of diabetes, provides an opportunity to study the autoantibodies of older-adult onset type 1 diabetes without inclusion of people with a high probability of having non-type 1 diabetes.

In this study, we used a type 1 diabetes genetic risk score (T1DGRS) in clinically suspected type 1 diabetes cases to confirm type 1 diabetes and aimed to evaluate the prevalence and pattern of islet autoantibodies (GADA, IA2A and ZnT8A) at diagnosis in type 1 diabetes occurring after 30 years of age.

## Method

### Study population

Our study population of clinically diagnosed type 1 diabetes was a part of a UK wide study (ADDRESS-2). The detailed study protocols have previously been published (25). Participants with a clinical diagnosis of type 1 diabetes were recruited within 6 months of type 1 diabetes diagnosis, insulin treated from diagnosis and aged over 4 years at recruitment. Type 1 diabetes was clinician-diagnosed and biomarker confirmation was not required prior to recruitment. DNA and serum samples were collected alongside clinical characteristics and symptoms at diagnosis. Participants recruited up to September 2017 were included in the present analysis. We studied those who self-reported as being White European (n=1866), the only ethnicity for which the T1DGRS risk score has been validated. In those aged 18 years or younger at recruitment, body mass index (BMI) was age adjusted to adult levels using UK-WHO (2007) reference data (26). We analysed participants by childhood onset diabetes (<18 years), young-adult onset diabetes (18 to 30 years) and older-adult onset diabetes (>30 years).

### Laboratory analysis

Analysis of the islet autoantibodies glutamate decarboxylase (GADA), islet antigen-2 (IA2A) and zinc transporter 8 (ZNT8A) were performed centrally using an established radiobinding assay (27, 28). Islet autoantibodies were considered positive as published elsewhere (4) and was in line with the majority of assays in the Standardization Program Workshop.

### Generation of a type 1 diabetes Genetic Risk score

We generated a type 1 diabetes genetic risk score (T1DGRS), a measure of an individual’s genetic susceptibility to type 1 diabetes, from 30 common genetic variants associated with type 1 diabetes as previously described (29, 30). SNPs were directly genotyped by LGC genomics, as previously described (29) and the score generated by summing the effective allele dosage of each variant multiplied by the natural log (ln) of the odds ratio (29, 30).

### Testing for monogenic forms of diabetes

The coding regions and 50 nucleotides of flanking intronic sequence of 26 known monogenic diabetes genes and the mitochondrial DNA mutation m.3234A>G were analyzed using the Agilent SureSelect custom capture library and an Illumina NetSeq 500 sequencing platform according to the methodology described by Ellard *et al* (31). Interpretation and classification of sequence variants was undertaken based on the American College of Medical Genetics and Genomics (ACMG) guidelines (32). Only variants classified as likely pathogenic (class 4) or pathogenic (class 5) were included in the study.

### Genetically confirming type 1 diabetes

Within each age group (<18y, 18-30y, >30y) we compared the distribution of the T1DGRS by four different categories based on the number of positive islet autoantibodies (0, 1, 2 and 3). We then compared the distribution of the T1DGRS in these categories to the distribution of T1DGRS from a large non-type 1 diabetes (type 2 diabetes) population from the Wellcome Trust case control consortium (WTCCC) (n=1924) (33). Of note in non-T1D, T1DGRS is comparable irrespective of the underlying cause of non-type 1 diabetes being type 2, monogenetic or secondary diabetes (29, 30). This enabled us to determine the likelihood of genetically consistent type 1 diabetes being present in each category. Previous studies have shown genetic susceptibility changes with increasing age of diagnosis of type 1 diabetes therefore T1DGRS was analysed separately within each age group (13, 34–36). The proportion of genetically consistent type 1 diabetes was calculated for each group of interest using the following formula as previously described (24).

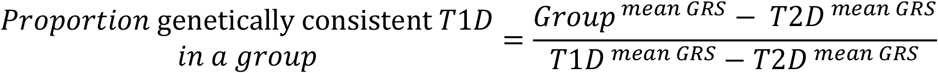

The mean T1DGRS of the group of interest (*Group*^*mean GRS*^) is evaluated relative to the mean T1DGRS of confirmed type 1 diabetes cases in the respective age groups (*T*1*D* ^*mean GRS*^) and the mean score of the references non-type 1 diabetes (WTCCC type 2 diabetes) population (*T*2*D* ^*mean GRS*^) in the above calculation (33).

### Statistical analysis

For comparisons made between groups separated by age and autoantibody status, Chi squared analysis was used for categorical characteristics and for continuous data we performed t tests for significance. One way ANNOVA was used to compare T1DGRS across three categories. All analyses were performed using Stata 16 (StataCorp LP, College Station, TX).

## Results

### Clinical characteristics of the study population

We included 1814 people with clinically diagnosed type 1 diabetes in the analysis after excluding 52 cases who were missing islet autoantibodies or T1DGRS (Supplementary Figure 1). The majority of cases were hospitalised at diagnosis (74%), had severe hyperglycaemia (mean HbA1c 87 mmol/mol) and were symptomatic (95% reporting polyuria or polydipsia, 85% weight loss, 41% presenting in diabetic ketoacidosis) (Supplementary Table 1). We analysed participants in three clinically relevant age groups: childhood onset (diagnosed aged <18 years, n=702), young-adult onset (diagnosed aged 18-30 years, n=524) and older-adult onset (diagnosed >30 years n=588). The clinical characteristics of these three groups are provided in Supplementary Table 1.

### In each age group all islet autoantibody positive cases have genetically consistent type 1 diabetes

Multiple positive islet autoantibody is highly specific for type 1 diabetes in those with a clinical suspicion of type 1 diabetes. Within each age group we first compared the T1DGRS of participants with one positive autoantibody to those with two or three positive islet autoantibodies. The T1DGRS was similar regardless of the number of positive islet autoantibodies in each age group [all p>0.1] (Supplementary Figure 2). The T1DGRS of islet autoantibody positive cases in each age group (0.277 (SD 0.026) <18 years, 0.271 (0.026) 18-30 years and 0.270 (0.027) >30years) was significantly higher than in people with type 2 diabetes (0.229, SD 0.034, n=1924) [all p<0.0001]. These data confirm that cases in all age groups with one, two or three positive islet autoantibodies have genetically consistent type 1 diabetes.

### T1DGRS of islet autoantibody negative adults was lower than genetically consistent type 1 diabetes cases and closer to the T1DGRS of type 2 diabetes cases

We next assessed the T1DGRS of people with negative islet autoantibodies and compared them to both the genetically consistent type 1 diabetes cases (all islet autoantibody positive cases) in the same age group and type 2 diabetes cases. We hypothesized that T1DGRS will be comparable if there is a similar proportion of genetically consistent type 1 diabetes between the groups, but T1DGRS will be lower if there is more non-type 1 diabetes in the autoantibody negative group. In children, the T1DGRS was similar between islet autoantibody negative and positive cases (mean 0.274 (SD 0.034) vs 0.277 (0.026)) [p=0.37] (Figure 1 and Supplementary Table 2). In adults, in contrast, the T1DGRS was substantially lower in islet autoantibody negative cases compared to positive cases (for young-adults (18-30y); mean T1DGRS 0.253 (0.039) vs 0.271 (0.026), for older-adults (>30y), 0.238 (0.034) vs 0.270 (0.027), p<0.0001 for both groups) (Figure 1, Table1 and Supplementary Table 3). The T1DGRS of negative islet autoantibody cases moved much closer to the T1DGRS of type 2 diabetes with increasing age of onset (mean 0.253 vs 0.229, difference 0.024 for the young-adult onset group and 0.238 vs 0.229, difference 0.09 for the older-adult onset group). This suggests a reducing presence of genetically consistent type 1 diabetes in older adults with clinically defined type 1 diabetes and negative islet autoantibodies.

**Table 1:**
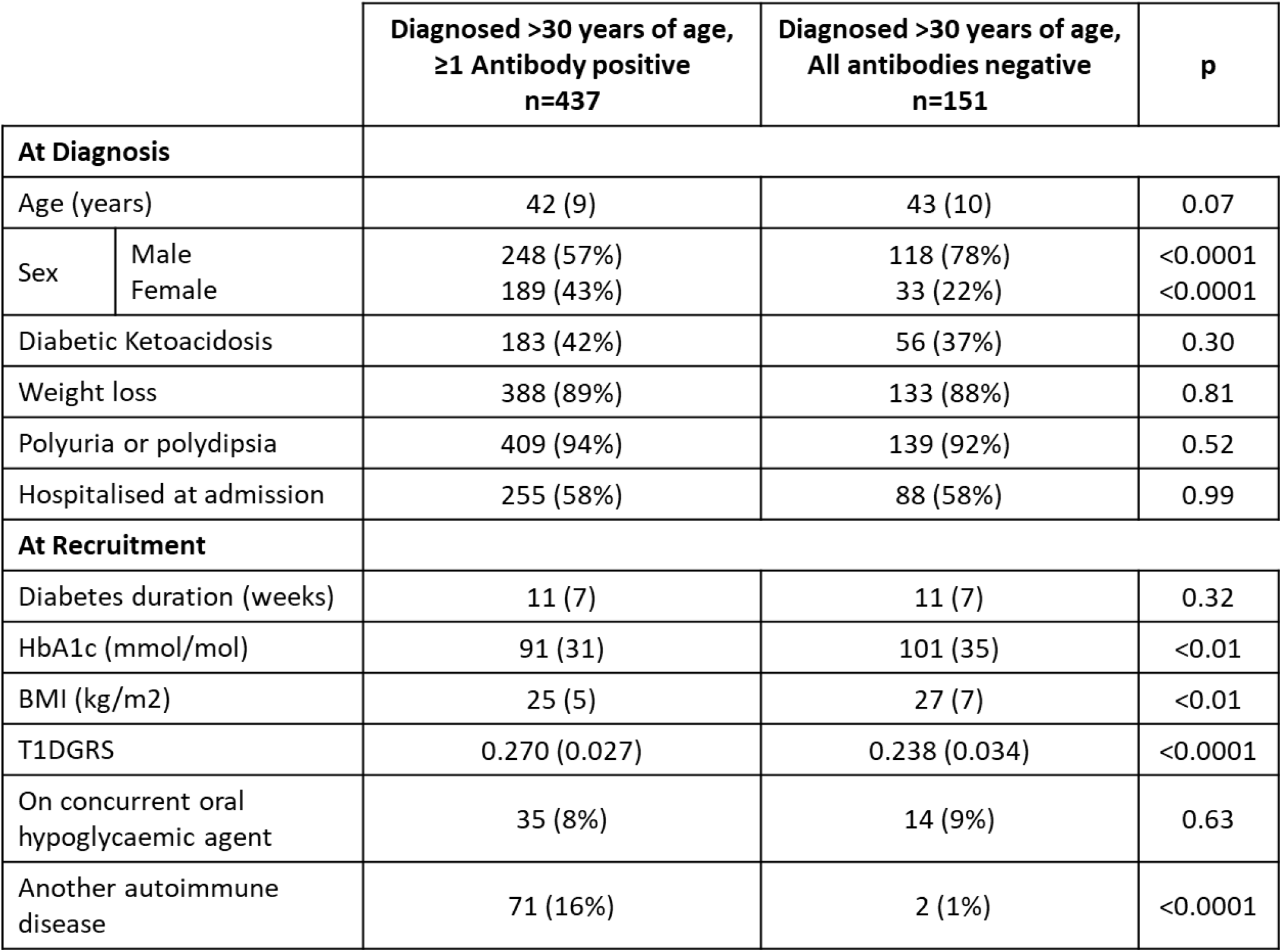
Clinical features of clinically diagnosed type 1 diabetes diagnosed >30 years of age split by islet autoantibody status. Data are mean (SD) or n (%). Missing variables have been treated as absent.

|  |  | Diagnosed >30 years of age,<br>≥1 Antibody positive<br>n=437 | Diagnosed >30 years of age,<br>All antibodies negative<br>n=151 | p |
| --- | --- | --- | --- | --- |
| **At Diagnosis** |  |  |  |  |
| Age (years) |  | 42 (9) | 43 (10) | 0.07 |
| Sex | Male | 248 (57%) | 118 (78%) | <0.0001 |
|  | Female | 189 (43%) | 33 (22%) | <0.0001 |
| Diabetic Ketoacidosis |  | 183 (42%) | 56 (37%) | 0.30 |
| Weight loss |  | 388 (89%) | 133 (88%) | 0.81 |
| Polyuria or polydipsia |  | 409 (94%) | 139 (92%) | 0.52 |
| Hospitalised at admission |  | 255 (58%) | 88 (58%) | 0.99 |
| **At Recruitment** |  |  |  |  |
| Diabetes duration (weeks) |  | 11 (7) | 11 (7) | 0.32 |
| HbA1c (mmol/mol) |  | 91 (31) | 101 (35) | <0.01 |
| BMI (kg/m<sup>2</sup>) |  | 25 (5) | 27 (7) | <0.01 |
| T1DGRS |  | 0.270 (0.027) | 0.238 (0.034) | <0.0001 |
| On concurrent oral hypoglycaemic agent |  | 35 (8%) | 14 (9%) | 0.63 |
| Another autoimmune disease |  | 71 (16%) | 2 (1%) | <0.0001 |

**Figure 1:**
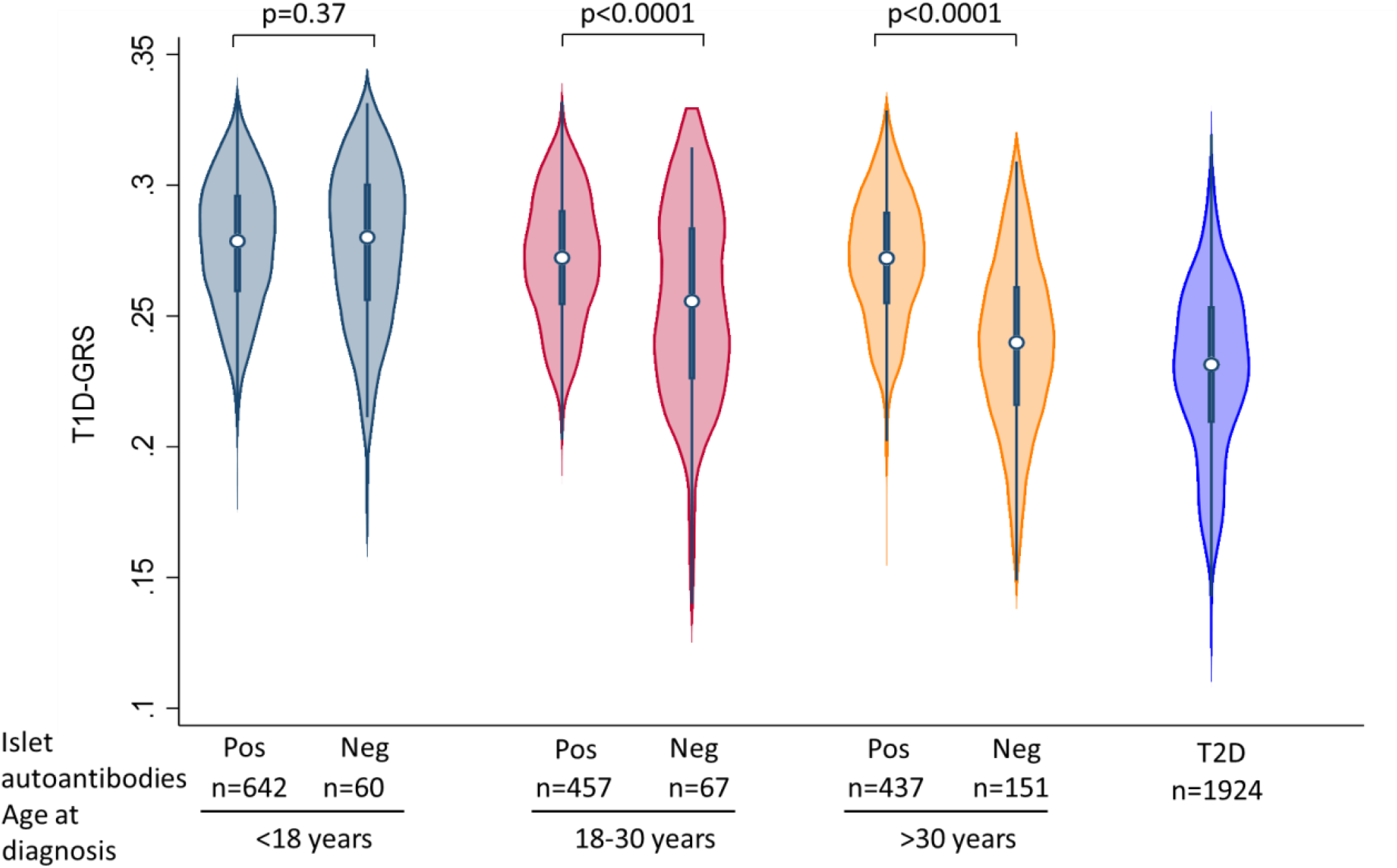
The type 1 diabetes genetic risk scores (T1DGRS) by age and islet autoantibody status in clinically diagnosed type 1 diabetes. The islet autoantibody positive group was positive for GADA, IA2A or ZnT8A whereas the islet autoantibody negative group was negative for all three islet autoantibodies. Type 2 diabetes individuals were from the Wellcome Trust Case control consortium (33).

### The T1DGRS of older adults with clinically diagnosed type 1 diabetes and negative islet autoantibodies is consistent with the majority having non-type 1 diabetes

We next sought to determine the proportion of islet autoantibody negative participants that had genetically consistent type 1 diabetes in the different age groups. Using the method described above and in our previous paper (37), we found that genetically consistent type 1 diabetes in 93% (n=56/60, 95% CI 84-98%) of autoantibody negative childhood cases, 55% (37/67, 95% CI 43-67%) of young-adult cases and only 23% of older-onset autoantibody negative cases (34/151, 95% CI 16-30%). This suggested that in autoantibody negative cases of clinically defined type 1 diabetes, 77% (95% CI 70-84%) of older-adults, 45% (95% CI 33-57%) of young-adults and 7% (95% CI 0-16%) of children had non-type 1 diabetes (misclassified as clinical type 1 diabetes). The overall misclassification was 1% (n=4/702, 95% CI 0-1%) in children, 6% (30/524, 95% CI 4-8%) for the young-adult group and 20% (117/588, 95% CI 17-23%) for the older-adult group.

Within islet autoantibody negative cases, genetic testing for all known causes of monogenic diabetes identified 5% (3/60, 95% CI 1-14%) of children, 8% (5/67, 95% CI 3-17%) of young-adults and 1% (2/151, 95% CI 0-5%) of older-adults with monogenic diabetes (Supplementary Figure 2). This data suggests that in children all misclassified patients had monogenic diabetes while in adults the majority had type 2 diabetes.

### Differences in clinical characteristics between islet autoantibody positive cases and islet autoantibody negative cases supports the presence of non-type 1 diabetes in adults

Despite all study participants having clinically diagnosed type 1 diabetes, the clinical characteristics of autoantibody negative cases were different to islet autoantibody positive cases supporting our results based on T1DGRS. Older-adults negative for all islet autoantibodies, compared to the islet autoantibody positive participants, were more likely to be male (78% vs 57%) and have a higher BMI (27 kg/m^2^ vs 25 kg/m^2^), and were far less likely to have a pre-existing autoimmune condition (1% vs 16%) [all p<0.01] (Table 1). Among young adults, those negative for all islet autoantibodies also had higher BMI (26 kg/m^2^ vs 24 kg/m^2^) compared to islet autoantibody positive individuals [p=0.01] (Supplementary Table 3). Islet autoantibody status had no meaningful impact on clinical characteristics in children in line with just 7% of islet autoantibody negative children having non-type 1 diabetes (Supplementary Table 2).

### Prevalence of positive islet autoantibody at diagnosis in genetically confirmed older-adult type 1 diabetes was the same as childhood-onset type 1 diabetes

We next assessed the islet autoantibody prevalence in all age groups in only genetically consistent type 1 diabetes cases. Within genetically consistent type 1 diabetes cases, older-adult onset cases, 93% (437/471, 95% CI 90-95%) had a positive islet autoantibody similar to 92% (457/494, 95% CI 90-95%) in young-adults and 92% (642/698, 95% CI 90-94%) in children [p=0.9] (Figure 2). This suggests no difference in prevalence of positive islet autoantibodies between age groups in contrast to when type 1 diabetes is defined clinically (Supplementary table 1).

**Figure 2:**
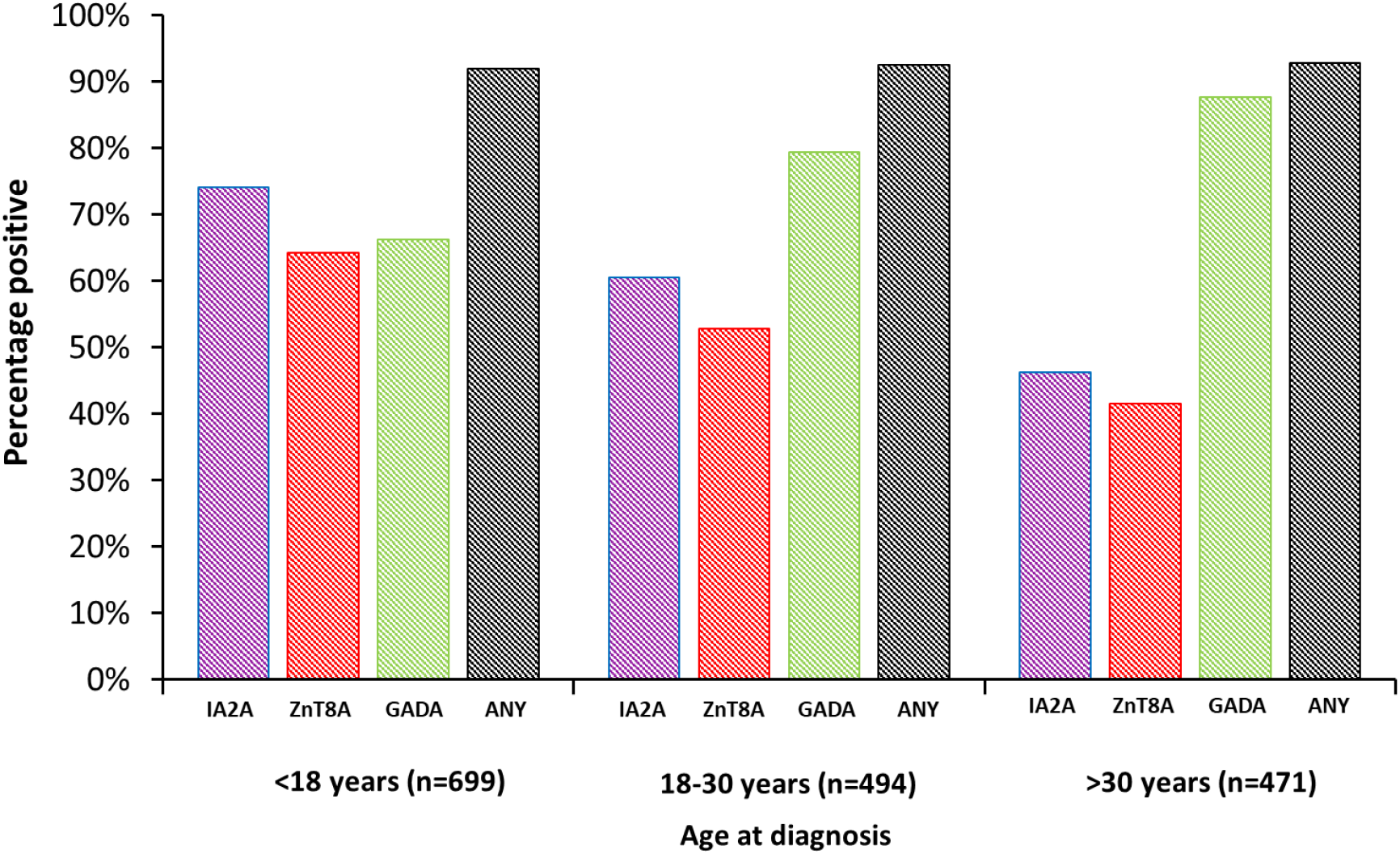
islet autoantibodies by age at diagnosis group in genetically consistent type 1 diabetes.

**Figure 3:**
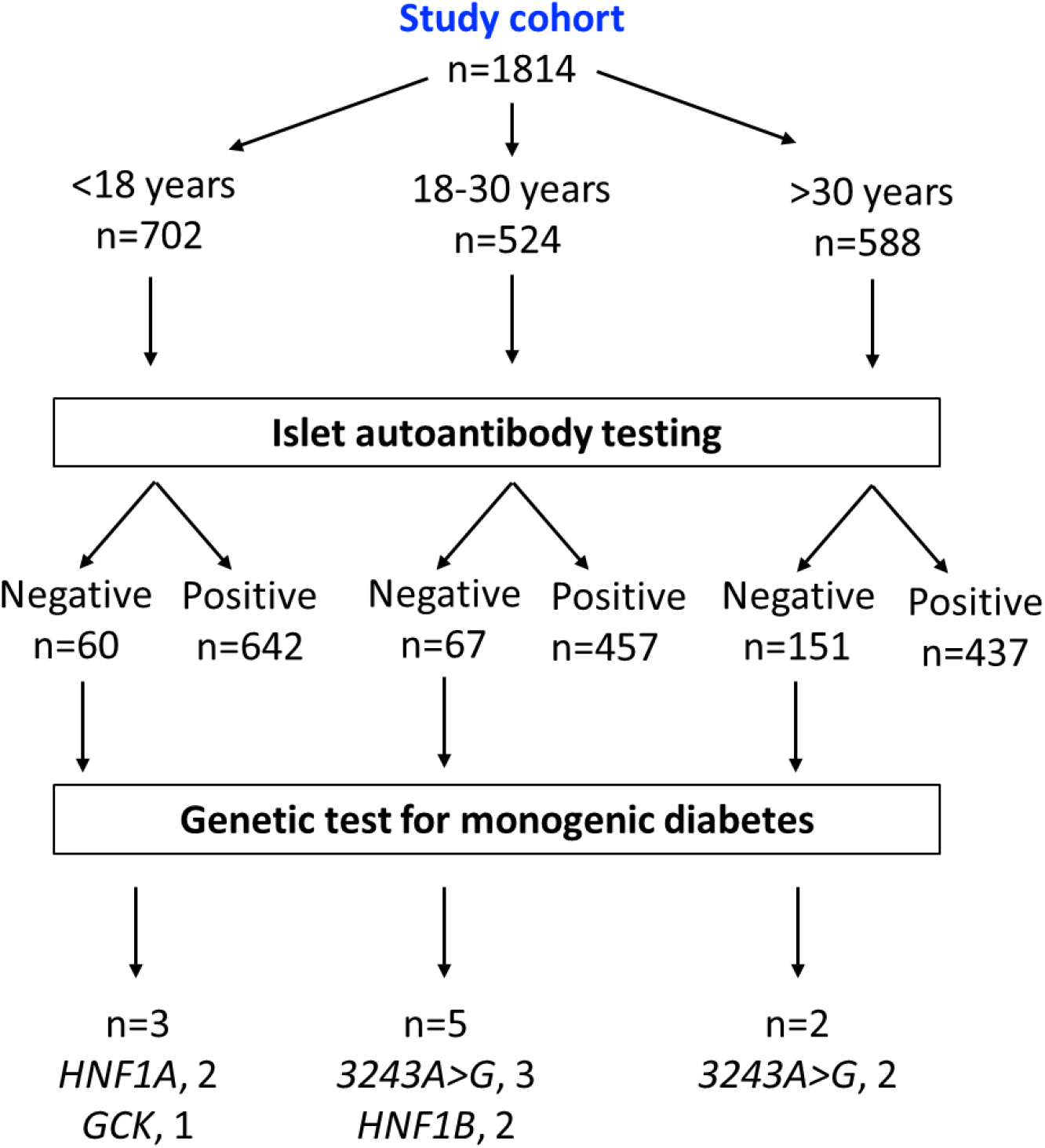
Flowchart showing results of mono-genetic analysis on islet autoantibody negative cases.

### Genetically consistent older-adult onset type 1 diabetes have different pattern of positive islet autoantibodies compared to childhood-onset type 1 diabetes with more single positive autoantibodies and predominance of GADA

The pattern of positive islet autoantibodies in the genetically consistent type 1 diabetes cases was different among the three age groups. Proportion with positive GADA increased with rising age of diagnosis: <18y 66% (463/698), 18-30y 79% (392/494), >30y 88% (413/471), [p<0.0001]. Conversely the proportion with positive IA2A and ZNT8A reduced with increasing age of diagnosis (for IA2A: 74% (517/698), 60% (299/494) and 46% (218/471) respectively; For ZNT8A: 64% (449/698), 53% (261/494) and 42% (196/471) respectively, [both p<0.0001]) (Figure 2 and Table 2). Older-adults were more likely to be single islet autoantibody positive (37%, 175/471) compared to young-adult and childhood onset cases (27%, 134/494 and 18%, 125/698 respectively) [p<0.0001] and less likely to have multiple positive islet autoantibodies (56%, 262/471) compared to young-adult and childhood onset cases (65% (323/494), and 74% (517/698) respectively) [p <0.0001] (Table 2). Almost all positive islet autoantibodies in those diagnosed ≥30 were captured by GADA (95%), with ZNT8A and IA2A together identifying additional genetically consistent type 1 diabetes cases in 5% of older-adults, 14% of young-adults and 28% of children (Figure 2) (Supplementary table 4).

**Table 2:**
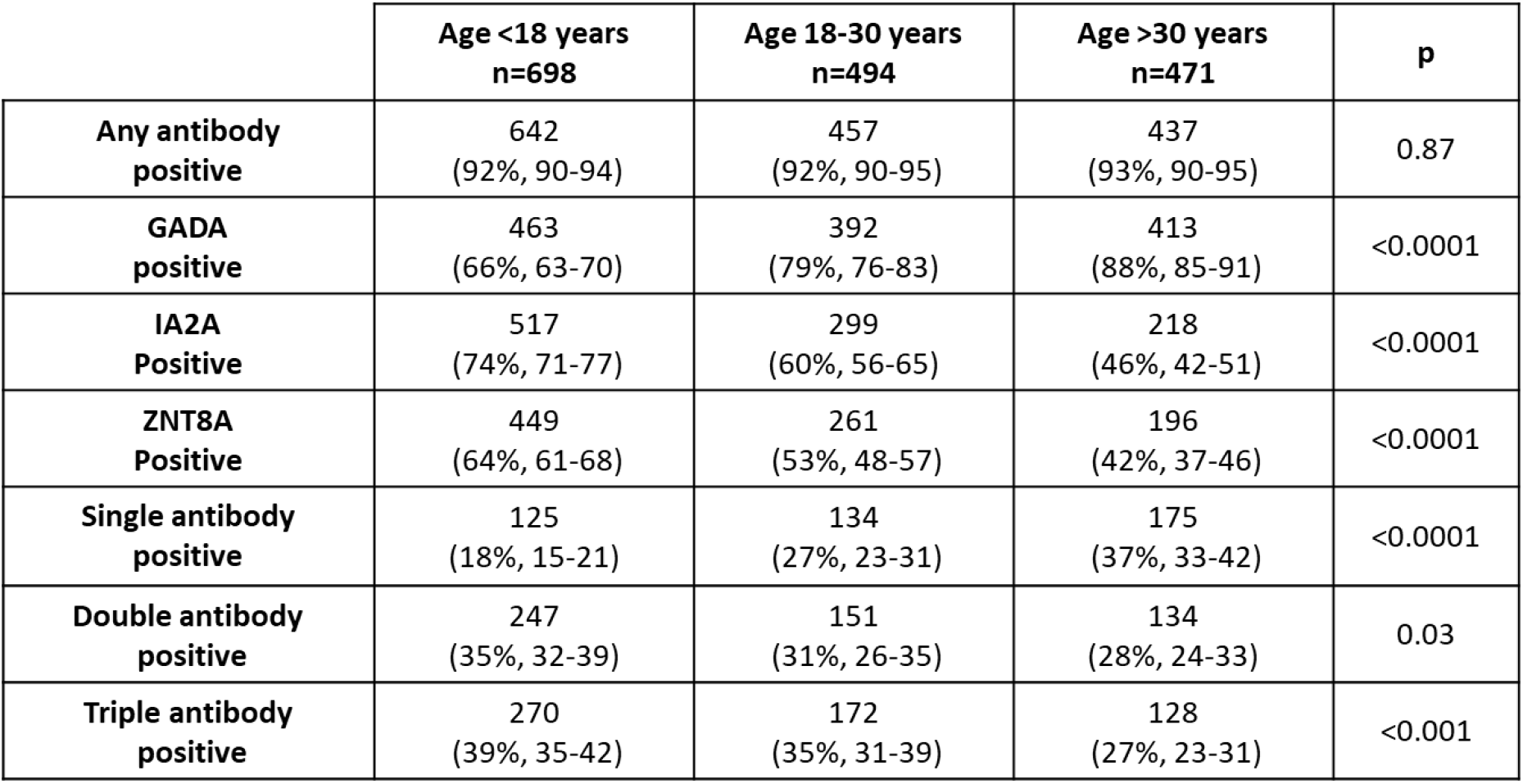
Pattern of positive islet autoantibodies in genetically consistent type 1 diabetes within three age at diagnosis groups. Data are n (%, 95% CI).

|  | **Age <18 years**<br>**n=698** | **Age 18-30 years**<br>**n=494** | **Age >30 years**<br>**n=471** | **p** |
| --- | --- | --- | --- | --- |
| **Any antibody**<br>**positive** | 642<br>(92%, 90-94) | 457<br>(92%, 90-95) | 437<br>(93%, 90-95) | 0.87 |
| **GADA**<br>**positive** | 463<br>(66%, 63-70) | 392<br>(79%, 76-83) | 413<br>(88%, 85-91) | <0.0001 |
| **IA2A**<br>**Positive** | 517<br>(74%, 71-77) | 299<br>(60%, 56-65) | 218<br>(46%, 42-51) | <0.0001 |
| **ZNT8A**<br>**Positive** | 449<br>(64%, 61-68) | 261<br>(53%, 48-57) | 196<br>(42%, 37-46) | <0.0001 |
| **Single antibody**<br>**positive** | 125<br>(18%, 15-21) | 134<br>(27%, 23-31) | 175<br>(37%, 33-42) | <0.0001 |
| **Double antibody**<br>**positive** | 247<br>(35%, 32-39) | 151<br>(31%, 26-35) | 134<br>(28%, 24-33) | 0.03 |
| **Triple antibody**<br>**positive** | 270<br>(39%, 35-42) | 172<br>(35%, 31-39) | 128<br>(27%, 23-31) | <0.001 |

## Discussion

This study shows novel insights into islet autoantibodies in type 1 diabetes at different ages of onset, using a methodology based on type 1 diabetes genetic predisposition. This methodology allows us to identify the proportion of genetically consistent type 1 diabetes within subgroups of clinically defined type 1 diabetes of different ages. In children the vast majority of antibody negative clinically diagnosed type 1 diabetes cases have genetically consistent type 1 diabetes with a minority having monogenic diabetes (<5%). In contrast in adults the majority of antibody negative clinically defined type 1 diabetes cases have type 2 diabetes and this is most marked in those diagnosed >30 years (77%). We show that the proportion of positive islet autoantibodies in genetically consistent older-adult onset type 1 diabetes (93%) is similar to genetically consistent type 1 diabetes presenting in children (92%) and young-adults (92%). We also describe the patterns of positive islet autoantibodies which are significantly different between age groups, with more older-adults having single positive islet autoantibodies and GADA predominating. Our data suggest that as in children, when adults have clinically suspected type 1 diabetes, positive islet autoantibodies confirm the diagnosis. However, contrary to in children, negative islet autoantibodies in adults (even where there is a high clinical suspicion of type 1 diabetes) suggests a high likelihood of non-type 1 diabetes. These findings have important clinical implications and support routine islet autoantibody testing in adults with clinically suspected type 1 diabetes.

We showed that, when the analysis is limited to genetically consistent type 1 diabetes, 93% of older-adult type 1 diabetes participants were positive for one of the three islet autoantibodies tested in this study which was similar to childhood-onset and young-adult onset cases.These finding contrasts with previous studies showing a reduction in positive islet autoantibodies (2-4, 13). The previous studies of older-adult onset type 1 diabetes used clinical diagnosis of type 1 diabetes alone and may have inadvertently included atypical non-type 1 diabetes cases leading to the apparent reduction in proportions of positive islet autoantibodies. Indeed, when we included all individuals with clinically diagnosed type 1 diabetes in our analysis, we also observed a reduction in positive islet autoantibody with increasing age of onset similar to published studies. This emphasises that the clinical presentation of type 2 diabetes can be severe and include features classically associated with type 1 diabetes. However, this will likely represent a very small fraction of the total type 2 diabetes population. A previous study which used insulin deficiency to define type 1 diabetes rather than clinical features also showed similar proportions of positive islet autoantibodies across age groups, although these were measured many years after diagnosis (12). Our study suggests that future studies to understand the biology of older-adult onset type 1 diabetes cases should not rely solely on clinical diagnosis alone and should include biomarkers such as islet autoantibodies or c-peptide as part of the definition.

We showed that despite having the same rate of positive islet antibodies, the overall pattern of islet autoantibodies was significantly different between older-adult and childhood-onset type 1 diabetes. Older-adults with genetically confirmed type 1 diabetes were more likely to have a single positive islet autoantibody with GADA the predominant antibody, present in 88% of older-adult cases. This finding is similar to the previous studies of older onset clinically diagnosed type 1 diabetes showing that when islet autoantibodies are present GADA predominates (4, 14). Coupled with the wide availability of GADA across the world, this makes GADA the most useful islet autoantibody to identify older-onset type 1 diabetes. In contrast in children and young adults our results confirm that all 3 autoantibodies should be measured as GADA will miss 28 % and 14% of the islet autoantibody positive cases respectively (18–30).

The T1DGRS of islet autoantibody positive cases was modestly reduced with increasing age of diagnosis, a finding that has been observed and reported previously (13, 34–36). We therefore compared the T1DGRS of autoantibody negative and positive cases within the same age group to ensure this did not impact our findings. The difference in patterns of islet autoantibodies and genetic predisposition with increasing age of genetically confirmed type 1 diabetes onset highlight the need for further detailed genetic, immunophenotyping and clinical studies to understand the biology of older-adult onset type 1 diabetes.

Our finding that when type 1 diabetes is diagnosed clinically, increasing age of onset associates with higher misclassification rates is remarkably consistent with studies defining diabetes type by C-peptide. In our study, T1DGRS was consistent with 77% of older-adult autoantibody negative clinically diagnosed type 1 diabetes participants having non-type 1 diabetes corresponding to 20% of this entire age group. As expected, this was lower in young-adults but still 45% of those negative for all islet autoantibodies and 6% of the entire age group. Our results are supported by another recent study which used C-peptide at over 3 years duration to define type 1 diabetes and found a similar rate of misclassification (20). Foteinopoulou et. al showed the presence of non-type 1 diabetes in 14% and 7% of all individuals who were clinically thought to have type 1 diabetes aged ≥30 years and 18-30 years respectively at diagnosis (20). All the misclassifications in their study were seen in antibody negative participants. This suggests the need for additional biomarkers or tools such as clinical diagnostic models at diagnosis in autoantibody negative cases to confirm type 1 diabetes (38). Our data suggest that T1DGRS may be additionally useful in this scenario and its utility has already been shown for discriminating type 1 diabetes from type 2 diabetes (29).

Our analysis was limited to White Europeans and therefore further research is required in different ethnicities, especially those where islet autoantibody proportions in clinically diagnosed type 1 diabetes are reported to be much lower (3). We did not have information on c-peptide at diagnosis which would have also helped to identify definite type 1 diabetes. However, whilst c-peptide confirms endogenous insulin deficiency in long-duration type 1 diabetes, it has limited utility close to diagnosis and thus unlikely to have changed the results of our study (21). We were only able to use a 30 variant T1DGRS in our study rather than recently published 67 variant T1DGRS score (39). This score only has modestly improved discriminatory power and therefore is unlikely to have significantly altered the results. Finally, this study was conducted in clinically defined type 1 diabetes cases and so the findings cannot be extended to all people presenting with diabetes as adults. This is highlighted by the comparable clinical characteristics between older adults with and without islet autoantibodies for example similar prevalence of diabetic ketoacidosis, despite a high rate of non-type 1 diabetes within antibody negative cases.

We believe that our results strongly suggest the need to change international clinical diabetes guidelines to measuring islet autoantibodies in all adults with suspected type 1 diabetes and not limiting it to those where there is diagnostic uncertainty (7, 8). We have shown the measurement of islet autoantibodies in clinical practice has different implications at different ages. In adults whilst c-peptide confirms endogenous insulin deficiency in long-duration type 1 diabetes, thought to have type 1 diabetes on clinical grounds, the measurement of islet autoantibodies can greatly assist this difficult clinical diagnosis. A positive islet autoantibody in all adults confirms the clinical diagnosis of type 1 diabetes. Conversely absence of all three islet autoantibody in adults makes the clinical diagnosis of type 1 diabetes unlikely, suggesting a high probability of non-type 1 diabetes (45% for 18-30y and 77% for >30). Therefore in adults with a clinical diagnosis of type 1 diabetes absence of islet autoantibodies should trigger an immediate re-assessment of the patient with serial evaluation of c-peptide. In older adults very few antibody positive cases will be missed if just GADA is measured. Conversely in children the benefit of testing autoantibodies is to help identify the small minority of patients with monogenic diabetes who will be negative for all islet autoantibodies. However it is important to appreciate that the vast majority (>90%) of antibody negative children will still have type 1 diabetes (1). If islet autoantibodies are measured in children, it is important to measure all islet autoantibodies.

## Conclusions

The proportion of positive islet autoantibodies in genetically consistent older-adult onset type 1 diabetes is comparable to childhood onset type 1 diabetes and almost entirely captured by GADA. Like children, positive islet autoantibodies in older adults with clinical suspicion of type 1 diabetes confirm the diagnosis. However, unlike children, negative islet autoantibodies in older adults with clinical suspicion of type 1 diabetes suggest a non-type 1 diabetes aetiology for the majority. Our study strongly suggests the routine measurement of islet autoantibodies in all adult cases with clinically suspected type 1 diabetes.

## Data Availability

ADDRESS 2 data access is available via a management committee

## Contributors

KAP, ATH, ADDRESS-2 study authors, and NJT designed the study. NJT, MNW and KP analysed the data. NJT wrote the first draft of the report. All authors reviewed the draft and contributed to the revision of the report.

## Acknowledgments

NJT is funded by a Wellcome Trust funded GW4 PhD. M.N.W. is supported by the Wellcome Trust Institutional Support Fund (WT097835MF). KAP is a Wellcome Trust fellow (219606/Z/19/Z) and the genetic risk score analysis in ADDRESS-2 was funded by Diabetes wellness Research foundation pump priming grant. ADDRESS-2 is co-funded by Diabetes UK (grant numbers 09-0003919, 15-005234 & 19/0006119) and the Juvenile Diabetes Research Foundation (grant numbers 9-2010-407 & 3-SRA-2015-35-A-N), supported by the NIHR Clinical Research Network and hosted by Imperial College London. S.M. is currently supported by a Future Leaders Mentorship award from the European Federation for the Study of Diabetes. NO is supported by the National Institute for Health Research (NIHR) Biomedical Research Centre based at Imperial College London. DJ is funded by the NIHR CRN. The views expressed are those of the authors and not necessarily those of the NHS, the NIHR, or the Department of Health

## Data Availability

ADDRESS 2 data access is available via a management committee (25).

## Ethics Statement

Ethics for the ADDRESS2 study was granted by the South Central – Berkshire NHS Research Ethics Committee on the 03/10/2010, ref: 10/H0505/85.

Ethics for monogenetic testing in the ADDRESS2 cohort was granted by the East of England – Essex Research Ethics Committee on the 14/7/2016, ref 16/EE/0306

## Supplementary

**Supplementary table 1:**
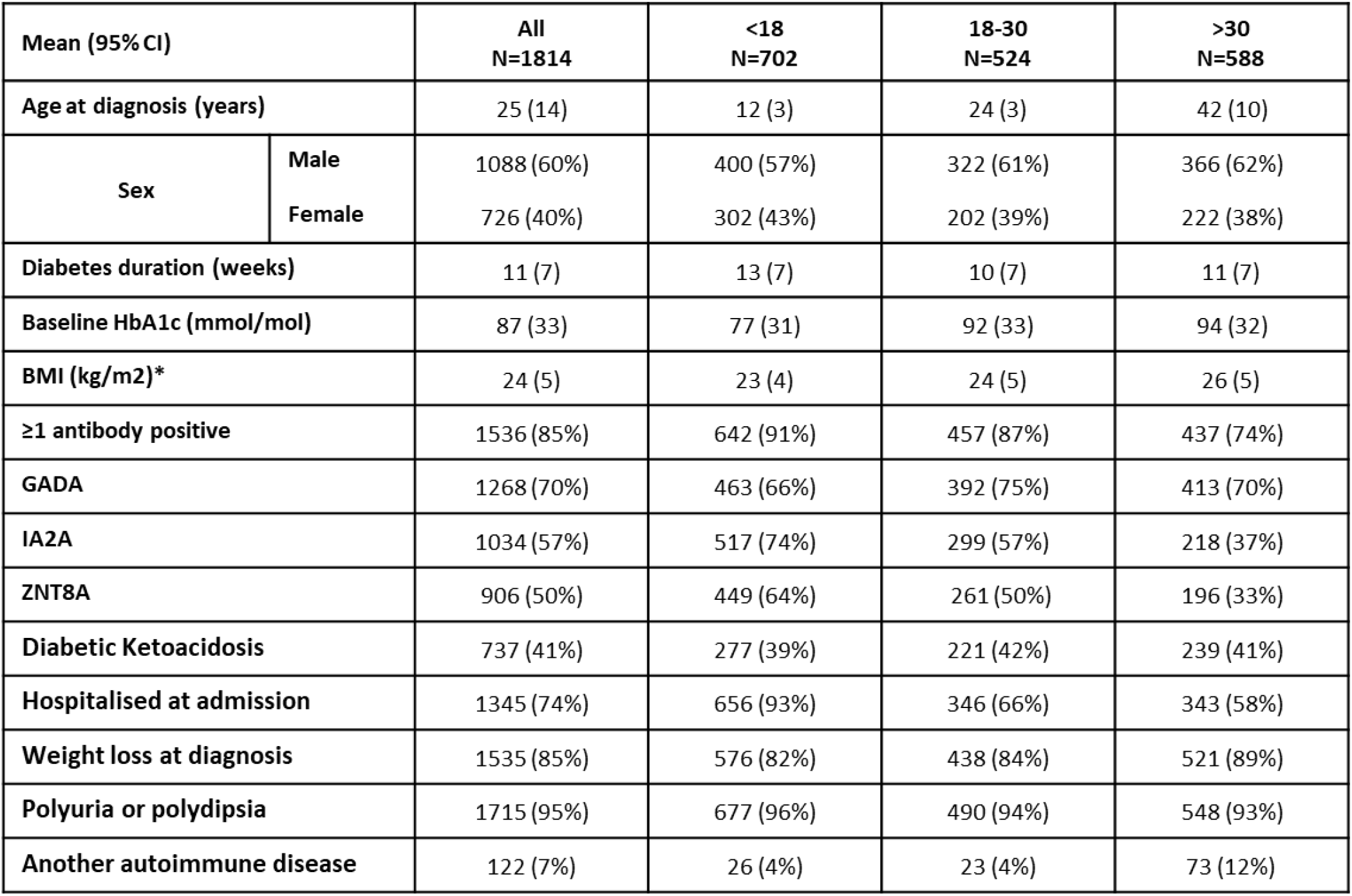
Clinically diagnosed type 1 diabetes baseline characteristics of whole cohort by age group. Data are mean (SD) or n (%). Missing variables have been treated as absent. * Body mass index (BMI) was age adjusted using WHO (2007) reference data (26)

**Supplementary table 2:**
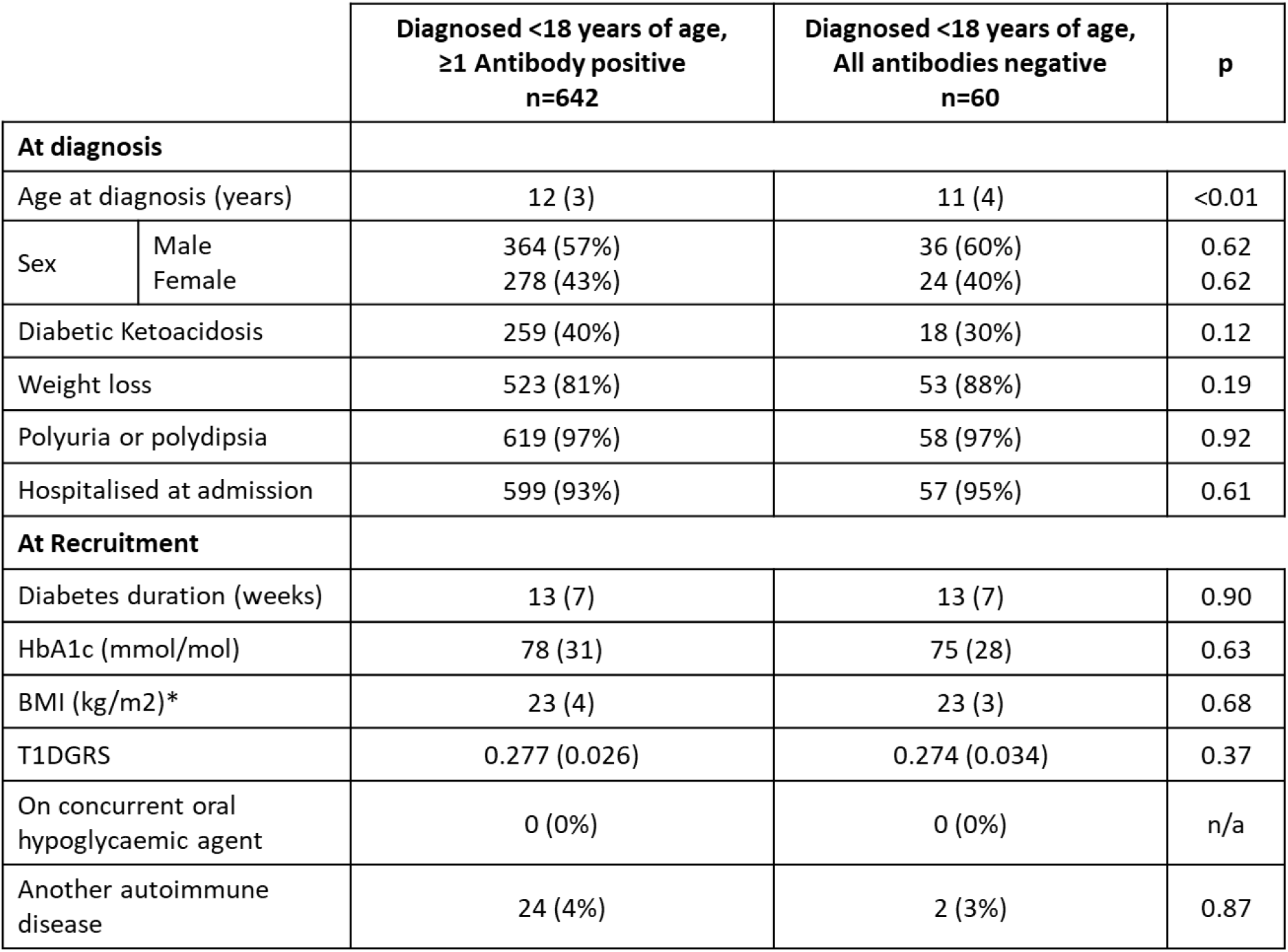
**Clinical features of clinically diagnosed type 1 diabetes diagnosed <18 years of age by islet autoantibody status.** Data are mean (SD) or n (%). Missing variables have been treated as absent. * Body mass index (BMI) was age adjusted using WHO (2007) reference data (26)

**Supplementary table 3:**
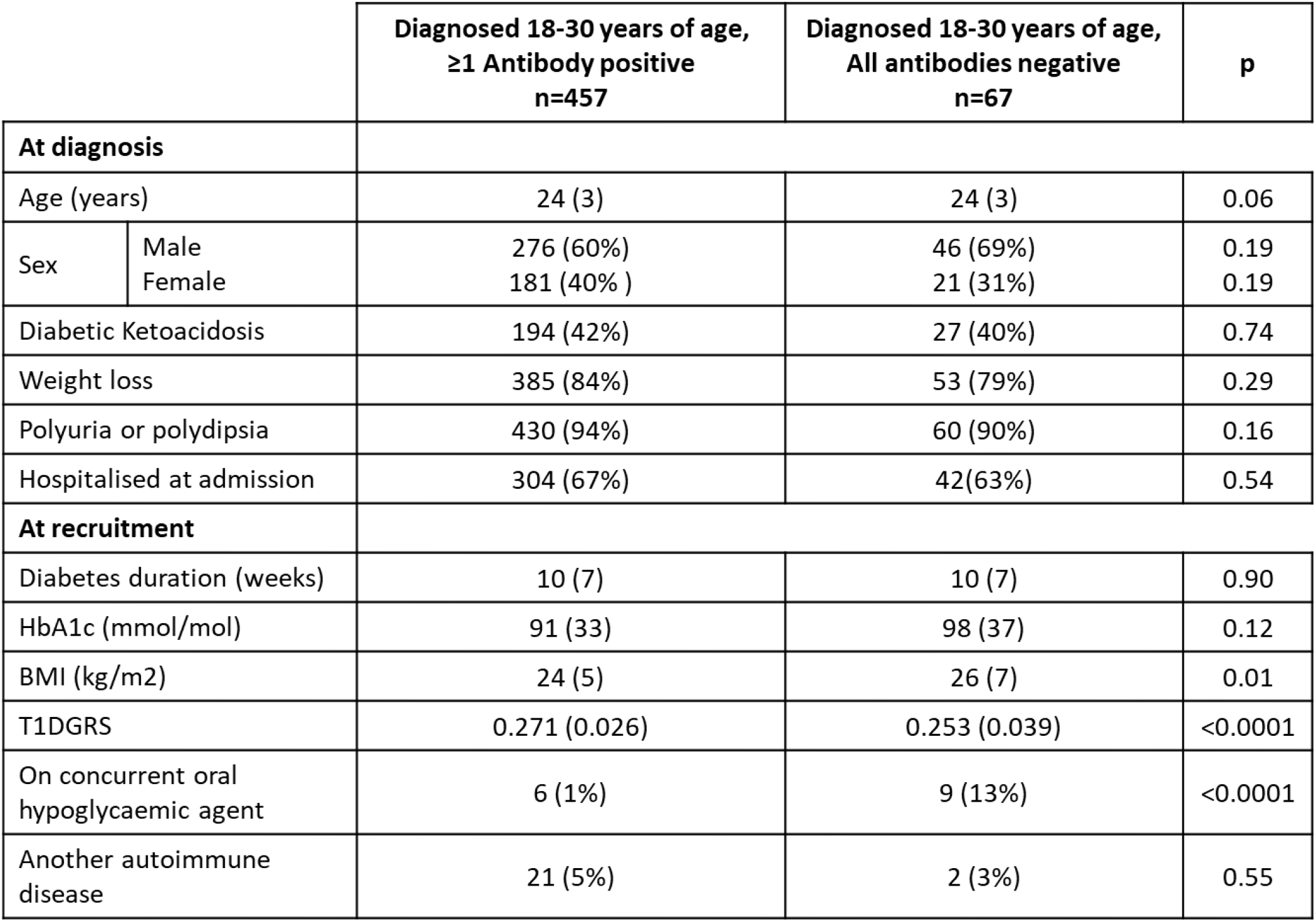
**Clinical features of clinically diagnosed type 1 diabetes diagnosed 18-30 years of age by islet autoantibody status.** Data are mean (SD) or n (%). Missing variables have been treated as absent.

**Supplementary table 4:**
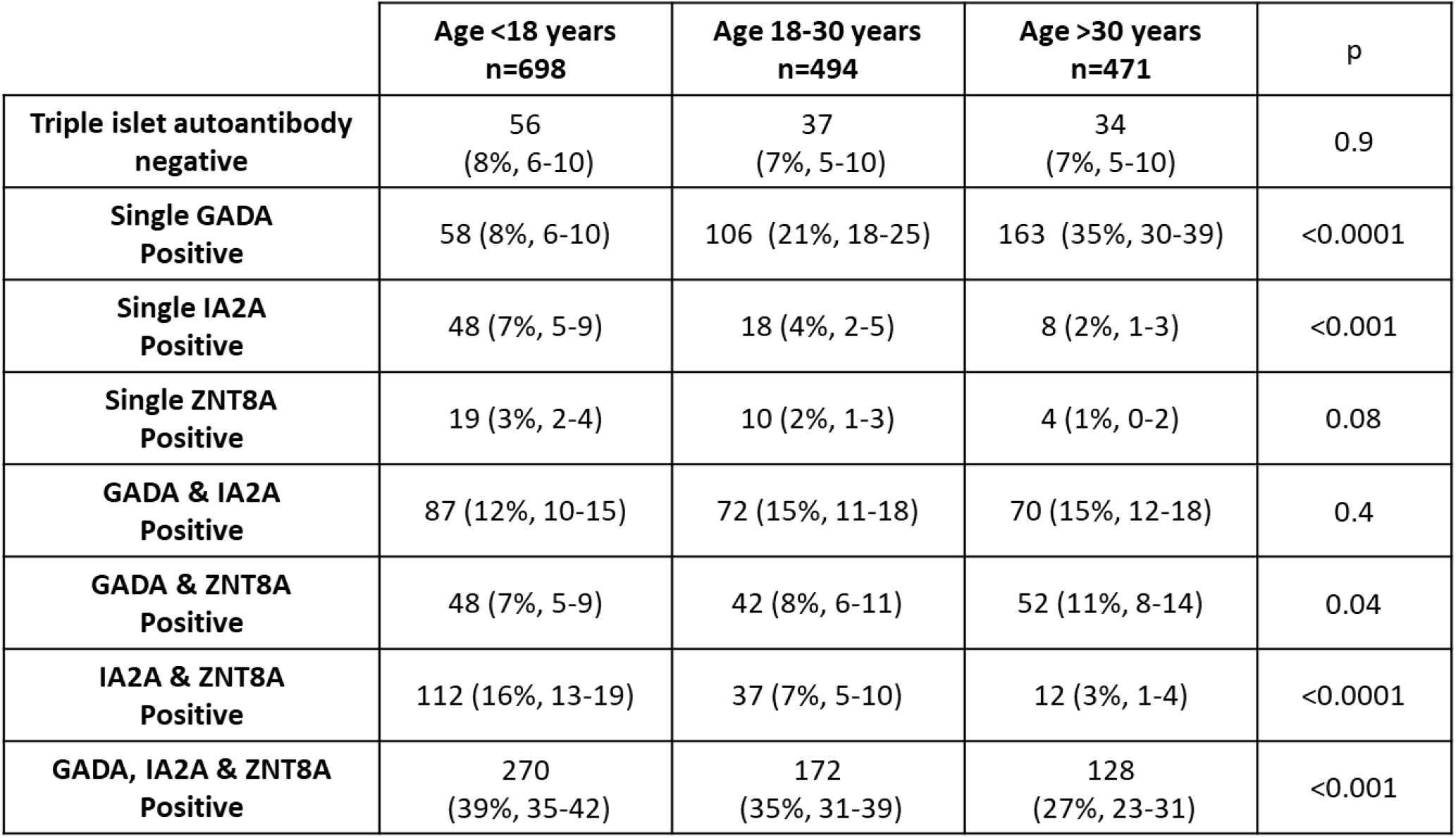
Pattern of islet autoantibody types in genetically defined type 1 diabetes by age group. Data are n (%, 95% CI)

**Supplementary figure 1:**
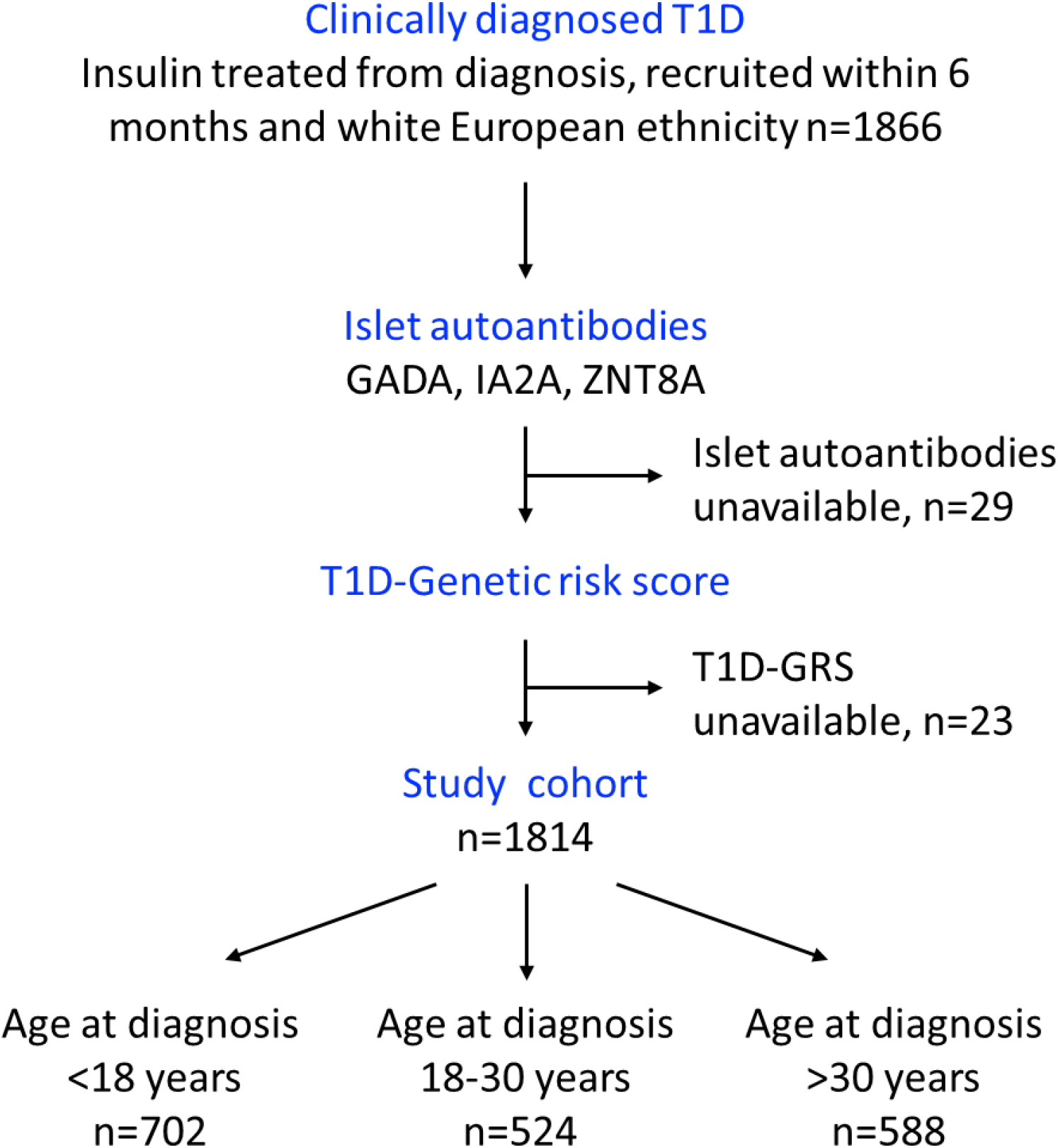
Flow diagram showing cohort selection and case exclusion.

**Supplementary figure 2:**
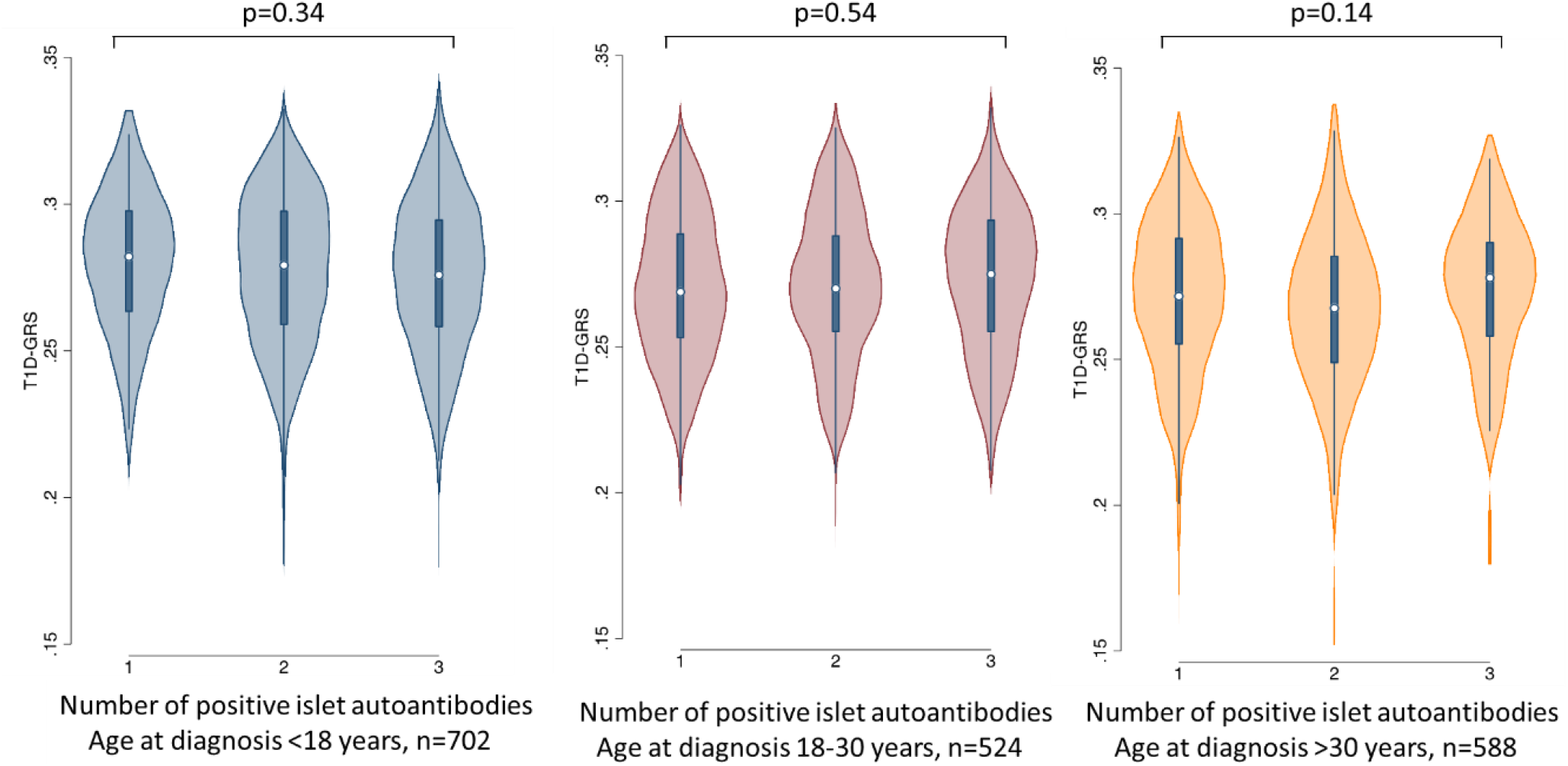
**Distribution of type 1 diabetes genetic risk score (T1DGRS) by number of positive islet autoantibodies in three age groups.**

