## Supplementary material for "The absence of islet autoantibodies in clinically diagnosed older-adult onset type 1 diabetes suggests an alternative pathology, advocating for routine testing in this age group"

**Supplementary table** 1: Clinically diagnosed type 1 diabetes baseline characteristics of whole cohort by age group. Data are mean (SD) or n (%). Missing variables have been treated as absent. * Body mass index (BMI) was age adjusted using WHO (2007) reference data (26)

| **Mean (95% CI)** | | **All N=1814** | **<18 N=702** | **18-30 N=524** | **>30 N=588** |
|---|---|---|---|---|---|
| **Age at diagnosis (years)** | | 25 (14) | 12 (3) | 24 (3) | 42 (10) |
| **Sex** | **Male** | 1088 (60%) | 400 (57%) | 322 (61%) | 366 (62%) |
| | **Female** | 726 (40%) | 302 (43%) | 202 (39%) | 222 (38%) |
| **Diabetes duration (weeks)** | | 11 (7) | 13 (7) | 10 (7) | 11 (7) |
| **Baseline HbA1c (mmol/mol)** | | 87 (33) | 77 (31) | 92 (33) | 94 (32) |
| **BMI (kg/m2)*** | | 24 (5) | 23 (4) | 24 (5) | 26 (5) |
| **≥1 antibody positive** | | 1536 (85%) | 642 (91%) | 457 (87%) | 437 (74%) |
| **GADA** | | 1268 (70%) | 463 (66%) | 392 (75%) | 413 (70%) |
| **IA2A** | | 1034 (57%) | 517 (74%) | 299 (57%) | 218 (37%) |
| **ZNT8A** | | 906 (50%) | 449 (64%) | 261 (50%) | 196 (33%) |
| **Diabetic Ketoacidosis** | | 737 (41%) | 277 (39%) | 221 (42%) | 239 (41%) |
| **Hospitalised at admission** | | 1345 (74%) | 656 (93%) | 346 (66%) | 343 (58%) |
| **Weight loss at diagnosis** | | 1535 (85%) | 576 (82%) | 438 (84%) | 521 (89%) |
| **Polyuria or polydipsia** | | 1715 (95%) | 677 (96%) | 490 (94%) | 548 (93%) |
| **Another autoimmune disease** | | 122 (7%) | 26 (4%) | 23 (4%) | 73 (12%) |

**Supplementary table 2: Clinical features of clinically diagnosed type 1 diabetes diagnosed <18 years of age by islet autoantibody status**. Data are mean (SD) or n (%). Missing variables have been treated as absent. * Body mass index (BMI) was age adjusted using WHO (2007) reference data (26)

| | | Diagnosed <18 years of age,<br>≥1 Antibody positive<br>n=642 | Diagnosed <18 years of age,<br>All antibodies negative<br>n=60 | p |
|---|---|---|---|---|
| **At diagnosis** | | | | |
| Age at diagnosis (years) | | 12 (3) | 11 (4) | <0.01 |
| Sex | Male | 364 (57%) | 36 (60%) | 0.62 |
| | Female | 278 (43%) | 24 (40%) | 0.62 |
| Diabetic Ketoacidosis | | 259 (40%) | 18 (30%) | 0.12 |
| Weight loss | | 523 (81%) | 53 (88%) | 0.19 |
| Polyuria or polydipsia | | 619 (97%) | 58 (97%) | 0.92 |
| Hospitalised at admission | | 599 (93%) | 57 (95%) | 0.61 |
| **At Recruitment** | | | | |
| Diabetes duration (weeks) | | 13 (7) | 13 (7) | 0.90 |
| HbA1c (mmol/mol) | | 78 (31) | 75 (28) | 0.63 |
| BMI (kg/m2)* | | 23 (4) | 23 (3) | 0.68 |
| T1DGRS | | 0.277 (0.026) | 0.274 (0.034) | 0.37 |
| On concurrent oral hypoglycaemic agent | | 0 (0%) | 0 (0%) | n/a |
| Another autoimmune disease | | 24 (4%) | 2 (3%) | 0.87 |

**Supplementary table 3: Clinical features of clinically diagnosed type 1 diabetes diagnosed 18-30 years of age by islet autoantibody status.** Data are mean (SD) or n (%). Missing variables have been treated as absent.

| | | **Diagnosed 18-30 years of age, ≥1 Antibody positive n=457** | **Diagnosed 18-30 years of age, All antibodies negative n=67** | **p** |
|---|---|---|---|---|
| **At diagnosis** | | | | |
| Age (years) | | 24 (3) | 24 (3) | 0.06 |
| Sex | Male | 276 (60%) | 46 (69%) | 0.19 |
| | Female | 181 (40% ) | 21 (31%) | 0.19 |
| Diabetic Ketoacidosis | | 194 (42%) | 27 (40%) | 0.74 |
| Weight loss | | 385 (84%) | 53 (79%) | 0.29 |
| Polyuria or polydipsia | | 430 (94%) | 60 (90%) | 0.16 |
| Hospitalised at admission | | 304 (67%) | 42(63%) | 0.54 |
| **At recruitment** | | | | |
| Diabetes duration (weeks) | | 10 (7) | 10 (7) | 0.90 |
| HbA1c (mmol/mol) | | 91 (33) | 98 (37) | 0.12 |
| BMI (kg/m2) | | 24 (5) | 26 (7) | 0.01 |
| T1DGRS | | 0.271 (0.026) | 0.253 (0.039) | <0.0001 |
| On concurrent oral hypoglycaemic agent | | 6 (1%) | 9 (13%) | <0.0001 |
| Another autoimmune disease | | 21 (5%) | 2 (3%) | 0.55 |

**Supplementary table 4:** Pattern of islet autoantibody types in genetically defined type 1 diabetes by age group. Data are n (%, 95% CI)

| | Age <18 years n=698 | Age 18-30 years n=494 | Age >30 years n=471 | p |
|---|---|---|---|---|
| **Triple islet autoantibody negative** | 56 (8%, 6-10) | 37 (7%, 5-10) | 34 (7%, 5-10) | 0.9 |
| **Single GADA Positive** | 58 (8%, 6-10) | 106 (21%, 18-25) | 163 (35%, 30-39) | <0.0001 |
| **Single IA2A Positive** | 48 (7%, 5-9) | 18 (4%, 2-5) | 8 (2%, 1-3) | <0.001 |
| **Single ZNT8A Positive** | 19 (3%, 2-4) | 10 (2%, 1-3) | 4 (1%, 0-2) | 0.08 |
| **GADA & IA2A Positive** | 87 (12%, 10-15) | 72 (15%, 11-18) | 70 (15%, 12-18) | 0.4 |
| **GADA & ZNT8A Positive** | 48 (7%, 5-9) | 42 (8%, 6-11) | 52 (11%, 8-14) | 0.04 |
| **IA2A & ZNT8A Positive** | 112 (16%, 13-19) | 37 (7%, 5-10) | 12 (3%, 1-4) | <0.0001 |
| **GADA, IA2A & ZNT8A Positive** | 270 (39%, 35-42) | 172 (35%, 31-39) | 128 (27%, 23-31) | <0.001 |

**Supplementary figure 1: Flow diagram showing cohort selection and case exclusion**

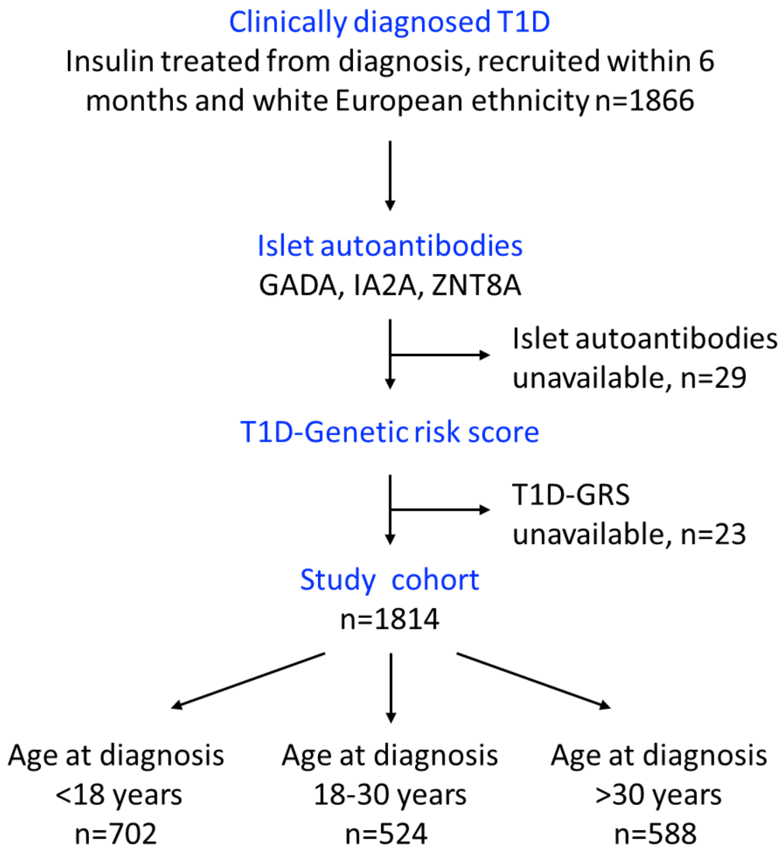

**Supplementary figure 2: Distribution of type 1 diabetes genetic risk score (T1DGRS) by number of positive islet autoantibodies in three age groups.**

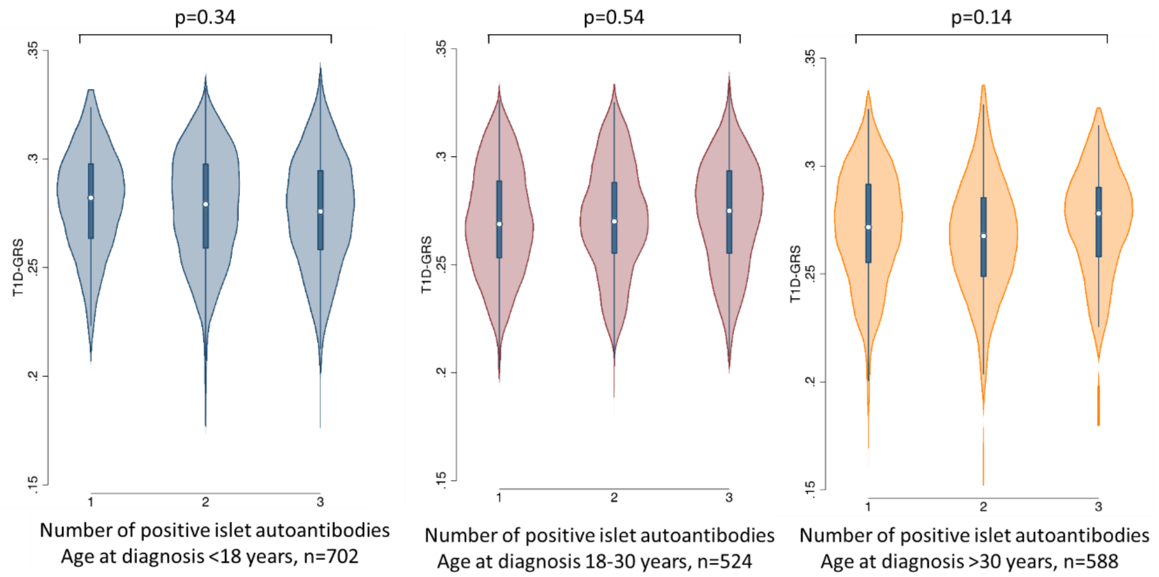

**Figure 3: Flowchart showing results of mono-genetic analysis on islet autoantibody negative cases**

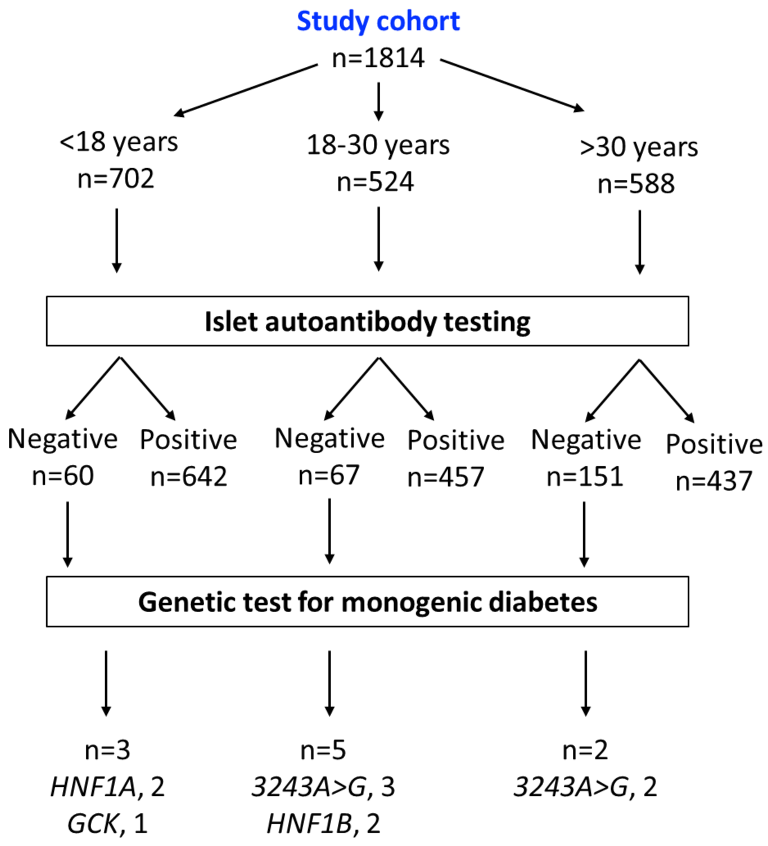
